# Understanding the prevalence of germline oncogenic biomarker variants across the Indian population

**DOI:** 10.1101/2023.10.20.23297298

**Authors:** Aastha Vatsyayan, Vinod Scaria

## Abstract

**Background:** Germline variants have traditionally been used for determining carrier status in heritable cancer families. However, studies are increasingly finding use of germline variants as therapeutic, diagnostic and prognostic biomarkers used to guide therapy decisions in cancer treatment.

**Objectives:** To study the prevalence of mutation specific oncogenic biomarkers in the Indian population and analyze their presence across disease cohorts

**Materials and Methods:** We annotate the IndiGen data obtained from whole genome sequencing of 1029 self-declared healthy Indian individuals with biomarker information obtained from the OncoKB knowledgebase, a repository of evidence-based information about somatic biomarkers and structural alterations in patient tumors.

**Results:** In our study we have discovered 34 biomarker variants of therapeutic actionability across 16 genes linked with 23 unique drugs or drug combinations in 23 unique types of cancer disease in the Indian population. In all, we have found 52 biomarker variants with 172 different biomarker types including therapeutic, resistance, diagnostic, and prognostic. We establish that 4.3% of the Indian population are carriers of therapeutic, and 2.13% are carriers of both diagnostic as well as prognostic germline biomarkers. Finally, we also establish the prevalence of 42 biomarker variants across the 23 genes in both AD and AR modes of inheritance in the Indian population.

## Introduction

Traditionally, germline variants in cancer have been studied mainly as predisposition biomarkers for surveillance and early detection of hereditary cancers. However, germline mutations can also determine response to both targeted, as well as traditional cancer therapy, including its toxicity. As Next-generation sequencing becomes increasingly accessible, several recent studies have highlighted the importance of studying germline variants as therapeutic markers. Several recent studies have observed germline mutations in cancer predisposition genes in sporadic cases with no family history of cancer^1^. Similarly, a pan-cancer analysis in 2017 demonstrated that approximately 17% of patients with advanced cancer had a pathogenic/likely pathogenic germline variant in a cancer susceptibility gene, with more than a quarter of these leading to consideration of change in therapy^2^.

The germline mutation burden is also observed to be high among pediatric cancer cases; A pan-cancer pediatric cohort analysis found that 6% of all patients carried causative germline variants, with 3% of all therapeutic biomarkers having a germline origin, with another pediatric study (CNS tumors) finding that 89% of participants harbored pharmacogenetic variants^2^. Additionally, pathogenic germline mutations have also been noted in a substantial number of sporadic pediatric cancer cases^1^. Thus application genomics is increasingly enabling the determination of the true burden of pathogenic germline mutations with therapeutic potential.

A common example of a biomarker-indicated drug is PARP inhibitors, approved for the treatment of metastatic breast cancer in germline BRCA mutation carriers^3^. Since germline mutations are clonal in nature (while a somatic mutation may be clonal as well as subclonal), a drug targeting a large clonal population has a higher chance of affecting a larger population of tumor cells, making it more important to study therapeutic implications of germline mutations^4^. Research efforts have also established variants as indicators of drug resistance/tolerance, efficacy, and toxicity (e.g. 5-fluorouracil related toxicity)^4^. Additionally, biomarkers can also offer diagnostic and prognostic value. Perhaps the most common example is the BCR-ABL1 fusion, which can used both for diagnosis of B-Lymphoblastic Leukemia/Lymphoma, and is also linked with a poor prognosis of the disease. Finally, the presence or absence of germline variants can guide surgical decisions (e.g. whether to perform mastectomy in women with BRCA1/2 mutations), as well as prophylactic decisions such as chemoprevention (e.g. prophylactic use of aspirin in carriers of Lynch syndrome)^4,5^.

In this study we utilize data from OncoKB^6^, a precision oncology knowledgebase that offers evidence-based information about somatic mutations and structural alterations in patient tumors, for annotating the IndiGen^7^ data, a database containing whole genome sequencing data of 1029 self-declared healthy Indian individuals from across 27 states. Using the resulting information, we calculate the prevalence of onco-biomarkers in the Indian population. We also query GUaRDIAN^8^, a nation-wide collaborative research initiative dedicated to the study of rare diseases to establish the presence of biomarkers across multiple cohorts. Finally, we compare our results with those obtained by cross referencing the MUSTARD^9^ resource, a database of mutation-specific therapies in cancer.

## Materials and Methods

### Biomarker Classification Based on Evidence-Based Clinical Actionability

We collected a list of 737 biomarker genes from the OncoKB database, and queried this list across the IndiGen compendium. We thus collected all germline variants reported across these biomarker genes. Next, using OncoKB API, we annotated this variant data with information regarding their biomarker status; OncoKB classifies variants into 4 main evidence based levels, each with several sub-levels of varying strength:

**Therapeutic (Levels 1-7):** Biomarkers predictive of response or resistance to a drug

**Diagnostic (Levels 1-2):** Biomarkers that facilitate diagnosis

**Prognostic (Levels 1-3):** Biomarkers that facilitate prognosis

**FDA (Levels 1-3):** Categorization of genetic tests by the FDA based on different levels of clinical significance

### Population Based Prevalence Analysis

We mapped global as well as subpopulation allele frequencies across all variants that had OncoKB annotations. We queried 14 global populations including IndiGen, GnomAD^10^, Human Genome Diversity Project (HGDP)^11^, TogoVar (Japanese population)^12^, the China Metabolic Analytics Project (ChinaMAP)^13^, Qatar^14^, the Singapore Sequencing Indian Project (SSIP)^15^, the Singapore Sequencing Malay Project (SSMP)^16^, 1000Genomes^17^, the Greater Middle East (GME) Variome Project^18^, Korea1K Variome^19^, Taiwan Biobank^20^, the Vietnamese Genetic Variation Database^21^, and the Iranome^22^. We performed the Fisher’s exact test across each of these populations and sub-populations with respect to the IndiGen population to determine whether the allele frequencies were significantly different in any population.

Next, based on the number of individuals who were carriers of any of the biomarkers, we established the percentage of biomarker carriers in the Indian populations. Finally, we calculated the incidence and prevalence of both Autosomal Dominant (AD) as well as Autosomal Recessive (AR) disorders linked with each of the genes containing the variants with OncoKB annotations in the IndiGen population.

### Disease Cohort Comparison

We queried all variants with OncoKB annotations across samples in the GUaRDIAN project^8^, a nation-wide collaborative research initiative that caters to rare diseases across multiple cohorts to determine cohort characteristics.

### Comparison with MUSTARD

We have previously compiled MUSTARD^9^, an exhaustive compendium of mutation-specific therapies in cancer. We queried both mutations as well as structural variants present in the database across OncoKB, and analyzed the variants that returned annotations.

## Results & Discussion

From our query of the IndiGen database, we found 17,00,332 variants across OncoKB biomarker genes. Upon annotating these, we obtained 2 Oncogenic, 330 Likely Oncogenic, 67 Likely Neutral, 32 Inconclusive and 16,99,900 Unknown variants. Of these 332 Oncogenic / Likely Oncogenic variants, 52 had highest level of annotations in any 4 of the evidence levels: a total of 34 variants had therapeutic biomarkers (28 Level 1, 3 Level 3A, 3 Level 4), 17 Diagnostic (11 Level Dx2, 6 Level Dx3), 11 Prognostic (5 Level Px1, 5 Level Px2, 1 Level Px3), and 34 had FDA levels (28 FDA Level 2, 6 FDA Level 3). Thus including all levels, 172 biomarker types (therapeutic, diagnostic, prognostic) were annotated across 27 unique genes, encompassing 23 unique drugs or drug combinations in 34 unique types of cancer disease in the Indian population. Complete details of the variants along with their annotations are shown in Supplementary Table 1.

### Variants Classified Based on Therapeutic Actionability

Of the therapeutic biomarkers, we found 34 biomarker variants across 16 unique genes (Figure 1). These variants included 56 Level 1 associations (defined as an FDA-recognized biomarkers predictive of response to an FDA-approved drug), 16 Level 2 associations (defined as standard care biomarkers recommended by the NCCN or other professional guidelines predictive of response to an FDA-approved drug), 23 Level 3A associations (defined as compelling clinical evidence supporting the biomarker as being predictive of response to a drug), and 9 Level 4 biomarkers (defined as compelling biological evidence supporting the biomarker as being predictive of response to a drug). In all, we discovered the presence of 118 biomarkers (therapeutic, diagnostic, and prognostic) in these 34 variants across 23 unique types of cancer, associated with 23 unique drugs.

**Figure 1:**
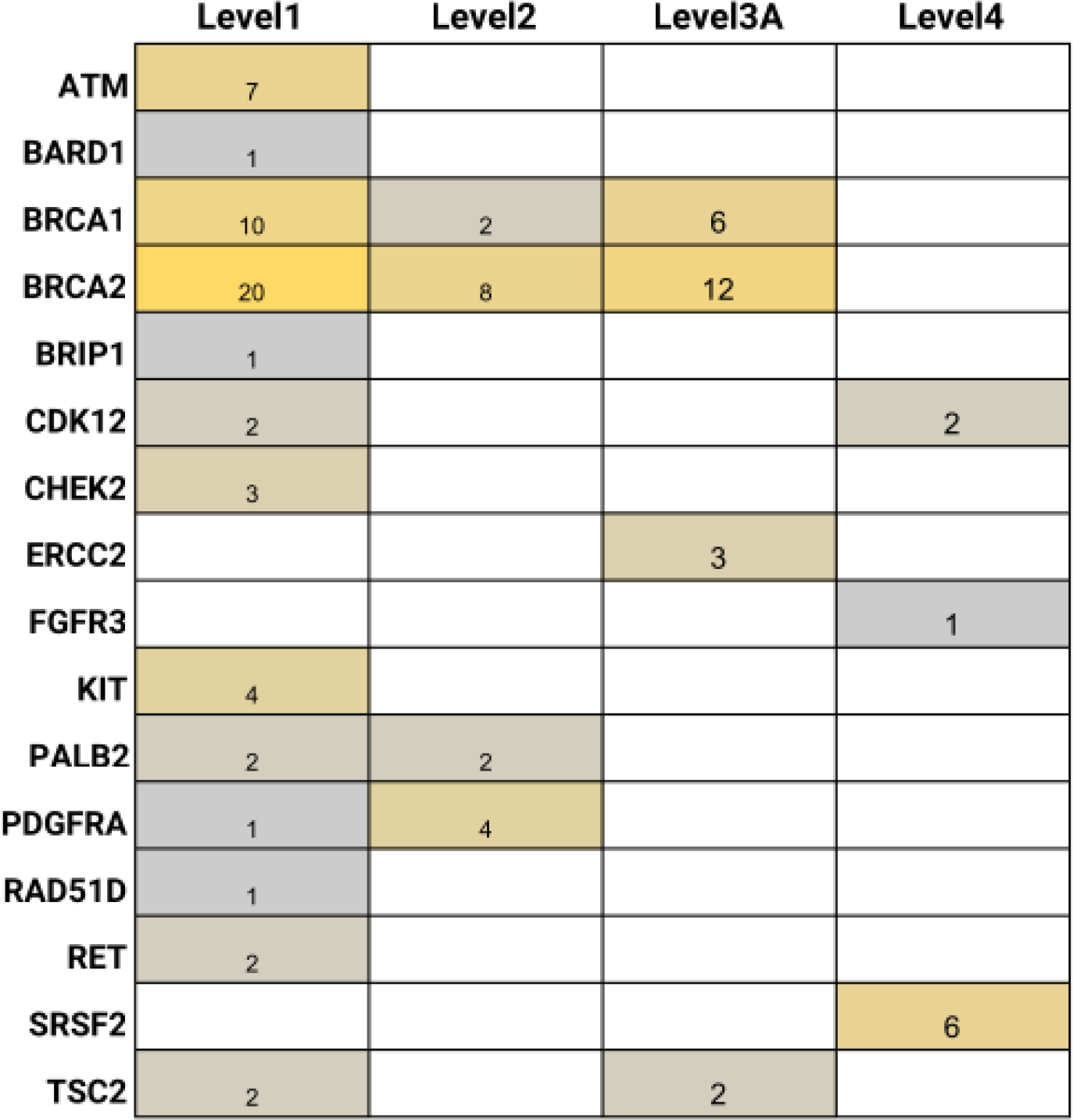
Figure describing all therapeutic biomarkers obtained across 34 IndiGen variants

Table 1 describes all the diseases and drugs associated with all 13 genes bearing Level 1 associations.

**Table 1:**
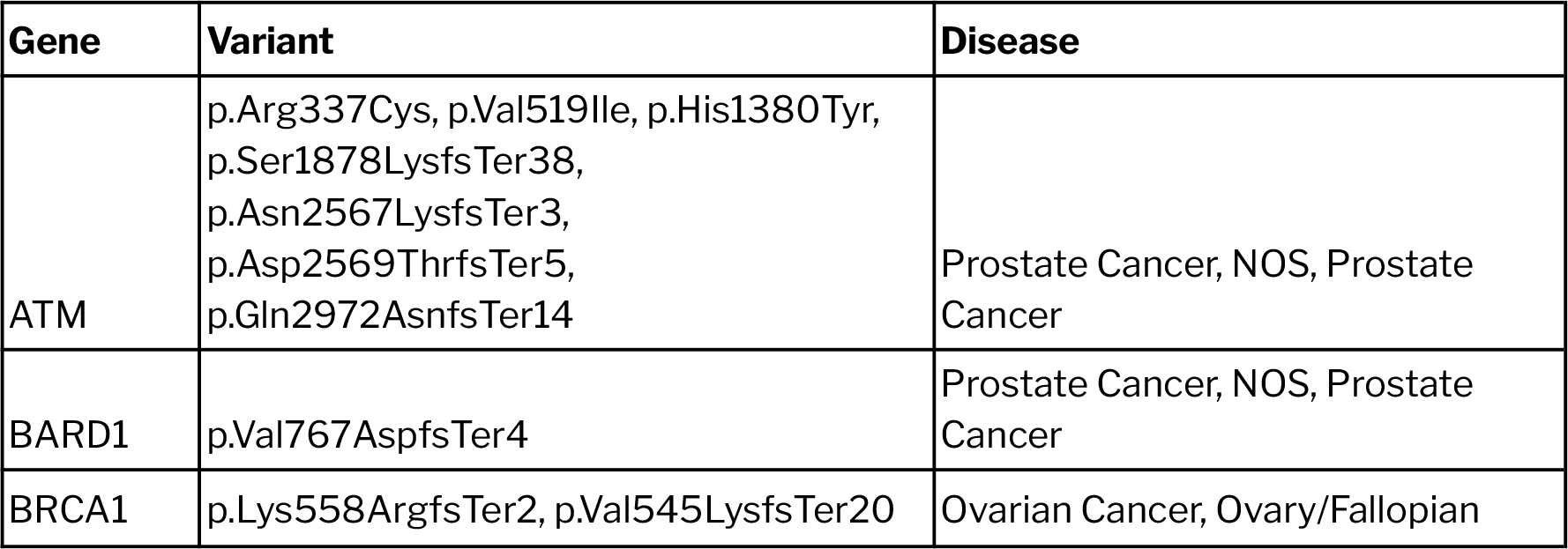
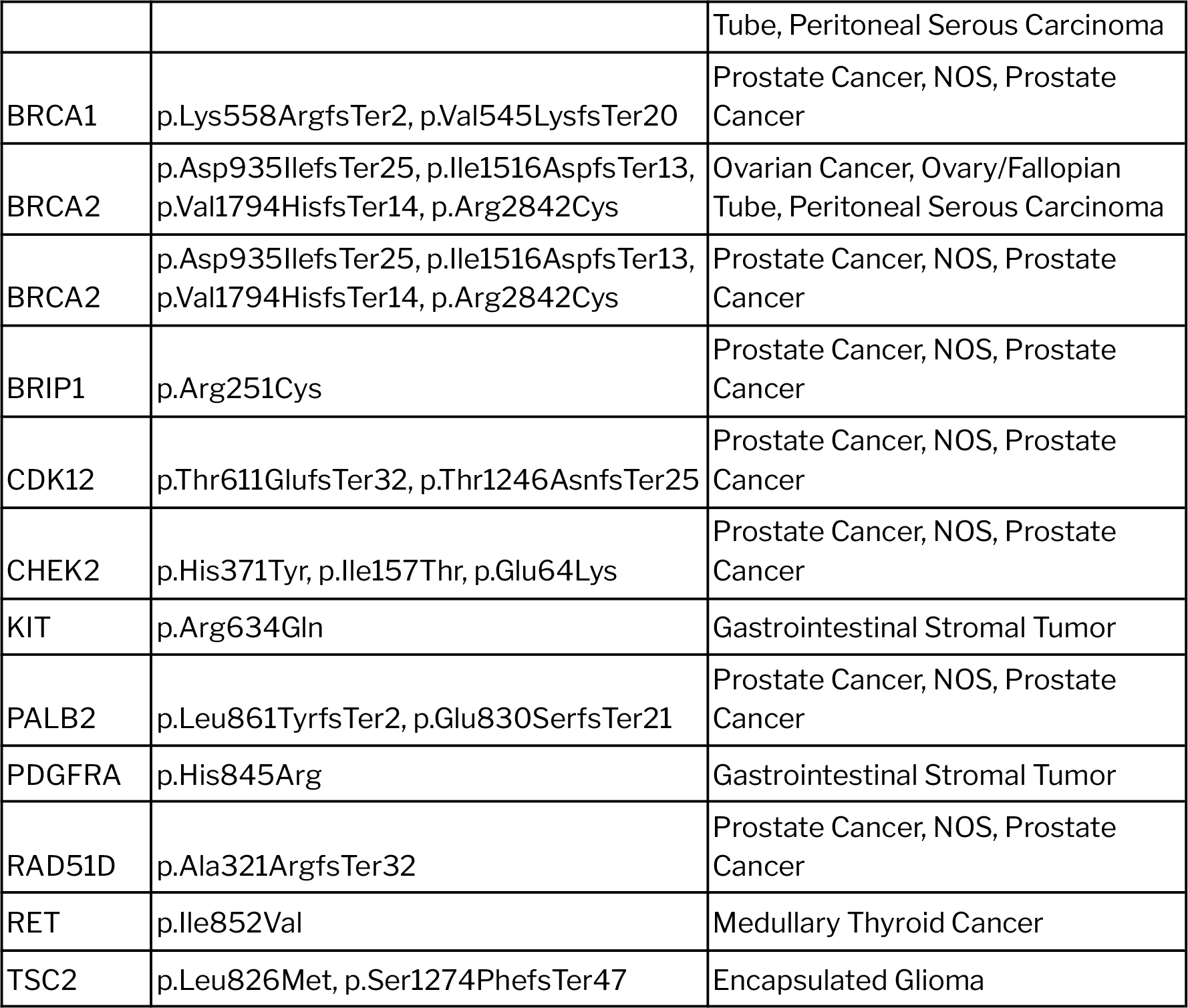
Table summarizing all Level1 therapeutic associations annotated across 13 genes in the IndiGen data.

### Population Based Prevalence Analysis

We found 8 variants from 7 genes that were significant across different populations at p-value < 0.01. Of these 8, the variant ATM/H1380Y was significant across all populations except Singapore, however, it was present at AF > 0.05 in the IndiGen (AF=0.064) and GnomAD Global (AF=0.066) populations. Additionally, it is also annotated as benign by ClinVar^23^. All other variants had AF < 0.05 across all populations. The plot depicting all non missing allele frequencies for the 52 variants is shown in Figure 2. The yellow circles mark statistically significant variants.

**Figure 2:**
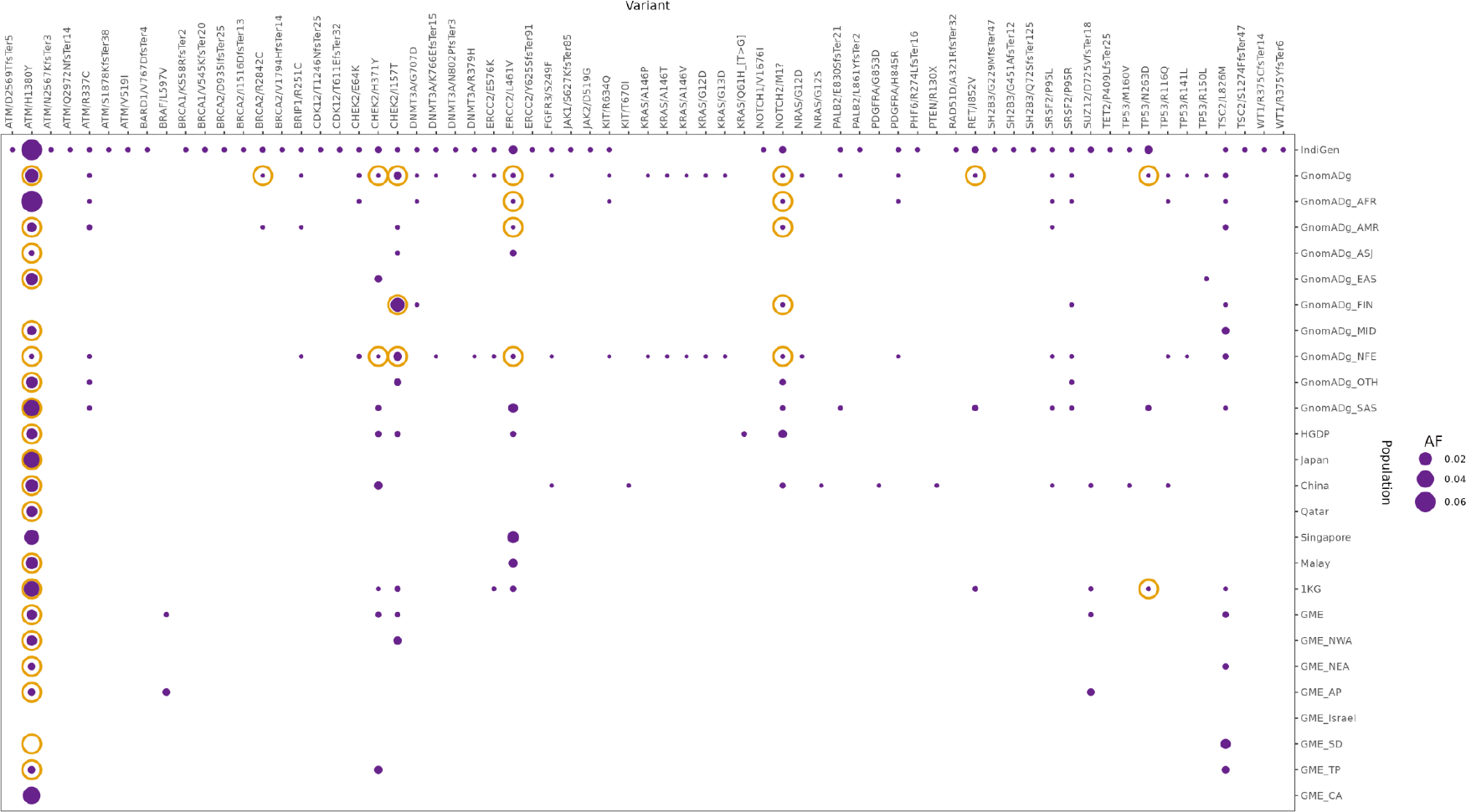
Plot depicting comparison between the allele frequencies of different populations bearing any of the 52 Oncogenic/Likely Oncogenic biomarker variants. The yellow circles highlight variants of statistical significance (p-value < 0.01)

Next, we calculated the prevalence of biomarkers across the Indian population using the IndiGen dataset. The variants with OncoKB annotations were linked with 27 genes. The ATM/H1380Y variant was removed from prevalence analyses. Additionally, 10 more variants had certain samples that did not pass QC, and calculations were made accordingly. Incidence and prevalence were calculated for each of the remaining 23 genes for both AD and AR modes of inheritance. Supplementary Table 2 shows these numbers along with the disorders associated with the genes according to OMIM^24^.

Notably, in the ERCC2 gene, 1 in about 85 individuals are expected to be carriers of pathogenic mutations linked with AD disease with known biomarkers. *ERCC2* is traditionally linked with AR inheritance of Xeroderma pigmentosum (OMIM). However, *ERCC2* mutations increase cisplatin sensitivity in bladder cancer cells, and can thus be used as a predictive biomarker in Muscle-Invasive Bladder Cancer^25^. Similarly, *TP53* is associated with an AD inheritance of Bone marrow failure syndrome among other cancers (OMIM), and is identified as a prognostic biomarker in several hematological malignancies. For example, *TP53*-mutated AML is associated with a lower likelihood of response to conventional chemotherapy and with poor outcomes, with a median overall survival of 4 to 6 months, and a 2-year overall survival rate of <10%^26^.

In all, 44 Therapeutic (24 Level 1, 7 Level 2, 8 Level 3A and 5 Level 4), 16 diagnostic and 16 prognostic biomarkers were observed. These mainly encompassed hematological malignancies (46%), breast and ovarian cancers (13%), prostate cancers (13%), and Gastrointestinal Stromal Tumors (11.8%). Other malignancies included pancreatic, Medullary thyroid cancer, Mesenchymal tumors, Uterine, Bladder, central nervous system, and all solid tumors.

#### Prevalence in India Population

Nearly 7% of the population (72 unique individuals) were found to be carriers of any one of the four evidence based biomarkers. Further, 4.3% (44 unique individuals) were carriers of therapeutic, and 2.13% (22 unique individuals) were carriers of both diagnostic as well as prognostic biomarkers.

### Disease Cohort Comparison

We discovered 10 unique variants across 365 samples from multiple cohorts in the GUaRDIAN data. These included all variants that were determined statistically significant except BRCA2/R2842C. Of these variants, two were annotated as diagnostic biomarkers: NOTCH2/M1? Which is a Diagnostic Level3 biomarker for Splenic Marginal Zone Lymphoma, and SUZ12/D725VfsTer18, a Diagnostic Level3 biomarker for Early T-Cell Precursor Lymphoblastic Leukemia. Of note, the SUZ12/D725VfsTer18 variant was present in several cohorts that represented diseases that increase the risk of hematologic malignancies, including Sjögren syndrome^27^, Mitochondrial Disorders^28^, Immunodeficiency^29^, Hyper IgE Syndromes^30^. The details of all 10 variants are shown in Supplementary Table 3.

### Comparison with MUSTARD

We queried the mutations and structural variants present across 168 unique genes from the MUSTARD database across OncoKB, and found 164 variants (66 Likely Oncogenic, 96 Oncogenic and 2 Resistance) with annotations across at least one of 4 evidence levels.

194 variants had Therapeutic evidence levels (63 Level 1, 62 Level 2, 8 Level 3A, 16 Level 4, 38 Level R1 and 7 Level R2), 92 had Diagnostic (9 Level Dx1, 55 Level Dx2, 28 Level Dx3), 46 had Prognostic (45 Level Px1, 1 Level Px2), and 149 had FDA level evidence (100 FDA Level 2, 49 FDA Level 3). The details of the variants along with their annotations are shown in Supplementary Table 4.

We next mapped all MUSTARD variants against the GUaRDIAN data; we found an overlap of 5 unique variants in 5 different samples. Two of the 5 variants had Level 1 biomarkers for breast cancer, and also belonged to the breast cancer cohort.

## Conclusions

In our study, we have calculated the prevalence of biomarkers implicated in cancer, that are of therapeutic, diagnostic and prognostic value in the Indian population. Some biomarkers such as ones for bladder, prostate and hematological malignancies are present in high numbers, and the information can be used to clinical advantage. However, only 23 of the 737 OncoKB genes queried had biomarkers in the Indian population. This could be because of differences in genetic makeup between different populations, and indicates a need for increased efforts to analyze biomarkers in variants prevalent in the Indian populations. Our comparison with the GUaRDIAN disease cohort data revealed concordance between two diagnostic biomarkers and their respective cohorts. 9 of the variants further lie in the top 10 most prevalent biomarker associated genes in the Indian population, showing further concordance between the two analyses. This illustrates the clinical advantage of studies like ours, where prevalence estimates can be used advantageously to guide biomarker research in a population specific manner.

Finally, our mapping of the MUSTARD database with the OncoKB knowledgebase yielded a greater overlap of variants than with the IndiGen data. Upon mapping MUSTARD with the GUaRIDIAN data, we found a similar concordance between the biomarkers and their respective cohorts, despite the small sample size.

## Data Availability

All data produced in the present work are contained in the manuscript.

## Acknowledgements

Authors acknowledge funding from the Council of Scientific and Industrial Research (CSIR) through CNP-0007 Grant. The funders had no role in the preparation of the manuscript or decision to publish.

## Author Contributions

VS conceptualized, designed and supervised the study. AV performed the analysis and complied the manuscript.

## Data Sharing And Availability

All data produced in the present work are contained in the manuscript.

## References

1. Kumar-Sinha, C. & Chinnaiyan, A. M. Precision oncology in the age of integrative genomics. Nat. Biotechnol. 36, 46–60 (2018).

2. Stadler, Z. K. et al. Therapeutic Implications of Germline Testing in Patients With Advanced Cancers. J. Clin. Oncol. 39, 2698–2709 (2021).

3. Tung, N. & Garber, J. E. PARP inhibition in breast cancer: progress made and future hopes. NPJ Breast Cancer 8, 47 (2022).

4. Thavaneswaran, S. et al. Therapeutic implications of germline genetic findings in cancer. Nat. Rev. Clin. Oncol. 16, 386–396 (2019).

5. Burn, J. et al. Cancer prevention with aspirin in hereditary colorectal cancer (Lynch syndrome), 10-year follow-up and registry-based 20-year data in the CAPP2 study: a double-blind, randomised, placebo-controlled trial. Lancet 395, 1855 (2020).

6. Chakravarty, D. et al. OncoKB: A Precision Oncology Knowledge Base. JCO Precis Oncol 2017, (2017).

7. Jain, A. et al. IndiGenomes: a comprehensive resource of genetic variants from over 1000 Indian genomes. Nucleic Acids Res. 49, D1225–D1232 (2021).

8. Sivasubbu, S. & Scaria, V. Genomics of rare genetic diseases—experiences from India. Hum. Genomics 13, 1–18 (2019).

9. Mittal, G. R I A., Vatsyayan, A., Pandhare, K. & Scaria, V. MUSTARD-a comprehensive resource of mutation-specific therapies in cancer. Database 2021, (2021).

10. Chen, S. et al. A genome-wide mutational constraint map quantified from variation in 76,156 human genomes. bioRxiv 2022.03.20.485034 (2022) doi:10.1101/2022.03.20.485034.

11. DeSalle, R. & Yudell, M. Welcome to the Genome: A User’s Guide to the Genetic Past, Present, and Future. (John Wiley & Sons, 2020).

12. Mitsuhashi, N. et al. TogoVar: A comprehensive Japanese genetic variation database. Human Genome Variation 9, 1–9 (2022).

13. Cao, Y. et al. The ChinaMAP analytics of deep whole genome sequences in 10,588 individuals. Cell Res. 30, 717–731 (2020).

14. Fakhro, K. A. et al. The Qatar genome: a population-specific tool for precision medicine in the Middle East. Hum Genome Var 3, 16016 (2016).

15. Wong, L.-P. et al. Insights into the genetic structure and diversity of 38 South Asian Indians from deep whole-genome sequencing. PLoS Genet. 10, e1004377 (2014).

16. Wong, L.-P. et al. Deep whole-genome sequencing of 100 southeast Asian Malays. Am. J. Hum. Genet. 92, 52–66 (2013).

17. A global reference for human genetic variation. Nature 526, 68–74 (2015).

18. Scott, E. M. et al. Characterization of Greater Middle Eastern genetic variation for enhanced disease gene discovery. Nat. Genet. 48, 1071–1076 (2016).

19. Jeon, S. et al. Korean Genome Project: 1094 Korean personal genomes with clinical information. Sci Adv 6, eaaz7835 (2020).

20. Wei, C.-Y. et al. Genetic profiles of 103,106 individuals in the Taiwan Biobank provide insights into the health and history of Han Chinese. NPJ Genom Med 6, 10 (2021).

21. Le, V. S. et al. A Vietnamese human genetic variation database. Hum. Mutat. 40, 1664–1675 (2019).

22. Fattahi, Z. et al. Iranome: A catalog of genomic variations in the Iranian population. Hum. Mutat. 40, 1968–1984 (2019).

23. Landrum, M. J. et al. ClinVar: improvements to accessing data. Nucleic Acids Res. 48, D835–D844 (2019).

24. Hamosh, A., Scott, A. F., Amberger, J. S., Bocchini, C. A. & McKusick, V. A. Online Mendelian Inheritance in Man (OMIM), a knowledgebase of human genes and genetic disorders. Nucleic Acids Res. 33, D514–7 (2005).

25. Li, Q. et al. ERCC2 Helicase Domain Mutations Confer Nucleotide Excision Repair Deficiency and Drive Cisplatin Sensitivity in Muscle-Invasive Bladder Cancer. Clin. Cancer Res. 25, (2019).

26. Short, N. J. et al. Prognostic and therapeutic impacts of mutant TP53 variant allelic frequency in newly diagnosed acute myeloid leukemia. Blood Adv 4, 5681–5689 (2020).

27. Lai, W.-S., Liu, F.-C., Wang, C.-H. & Chen, H.-C. Unusual cancer in primary Sjögren syndrome. Can. Fam. Physician 60, 912–915 (2014).

28. Sachdeva, A. et al. Association of leukemia and mitochondrial diseases-A review. J Family Med Prim Care 8, 3120–3124 (2019).

29. Raje, N., Snyder, B. L., Hill, D. A., Streicher, J. L. & Sullivan, K. E. Severe immunodeficiency associated with acute lymphoblastic leukemia and its treatment. Ann. Allergy Asthma Immunol. 120, 537–538.e1 (2018).

30. Freeman, A. F. & Holland, S. M. The hyper-IgE syndromes. Immunol. Allergy Clin. North Am. 28, 277–91, viii (2008).

